# Novel autoantibody targets identified in patients with autoimmune hepatitis (AIH) by PhIP-Seq reveals pathogenic insights

**DOI:** 10.1101/2023.06.12.23291297

**Authors:** Arielle Klepper, James Asaki, Andrew F. Kung, Sara E. Vazquez, Aaron Bodansky, Anthea Mitchell, Sabrina A. Mann, Kelsey Zorn, Isaac Avila-Vargas, Swathi Kari, Melawit Tekeste, Javier Castro, Briton Lee, Maria Duarte, Mandana Khalili, Monica Yang, Paul Wolters, Jennifer Price, Emily Perito, Sandy Feng, Jacquelyn J. Maher, Jennifer C. Lai, Christina Weiler-Normann, Ansgar W. Lohse, Joseph DeRisi, Michele Tana

## Abstract

**Background and Aims:** Autoimmune hepatitis (AIH) is a severe disease characterized by elevated immunoglobin levels. However, the role of autoantibodies in the pathophysiology of AIH remains uncertain.

**Methods:** Phage Immunoprecipitation-Sequencing (PhIP-seq) was employed to identify autoantibodies in the serum of patients with AIH (*n* = 115), compared to patients with other liver diseases (metabolic associated steatotic liver disease (MASH) *n* = 178, primary biliary cholangitis (PBC), *n* = 26, or healthy controls, *n* = 94).

**Results:** Logistic regression using PhIP-seq enriched peptides as inputs yielded a classification AUC of 0.81, indicating the presence of a predictive humoral immune signature for AIH. Embedded within this signature were disease relevant targets, including SLA/LP, the target of a well-recognized autoantibody in AIH, disco interacting protein 2 homolog A (DIP2A), and the relaxin family peptide receptor 1 (RXFP1). The autoreactive fragment of DIP2A was a 9-amino acid stretch nearly identical to the U27 protein of human herpes virus 6 (HHV-6). Fine mapping of this epitope suggests the HHV-6 U27 sequence is preferentially enriched relative to the corresponding DIP2A sequence. Antibodies against RXFP1, a receptor involved in anti-fibrotic signaling, were also highly specific to AIH. The enriched peptides are within a motif adjacent to the receptor binding domain, required for signaling and serum from AIH patients positive for anti-RFXP1 antibody was able to significantly inhibit relaxin-2 singling. Depletion of IgG from anti-RXFP1 positive serum abrogated this effect.

**Conclusions:** These data provide evidence for a novel serological profile in AIH, including a possible functional role for anti-RXFP1, and antibodies that cross react with HHV6 U27 protein.

## Introduction

Autoimmune hepatitis (AIH) is a chronic, severe liver disease identified in the 1950s, affecting all ages, rising in incidence^1^, and disproportionately impacting people of color^2,3^. Treatment of AIH frequently requires lifelong therapy with immunosuppressive medications, with multiple morbid side effects. Despite the longstanding clinical burden of AIH, little is known about the etiopathogenesis of disease. Clinically, AIH onset can be marked by an episode of acute hepatitis. Initial work-up includes assessment of total IgG levels (commonly elevated in patients with AIH), as well as evaluation for characteristic autoantibodies, particularly anti-nuclear antibodies (ANA), anti-smooth muscle antibodies (SMA), and liver-kidney microsomes type 1 (anti-LKM-1), none of which are specific to AIH or to the liver itself, as well as anti-soluble liver antigen and liver pancreas (SLA/LP), a liver antigen highly specific to AIH, present in up to 20% of AIH patients^4–6^. Determination of this serologic profile is essential for diagnosis and to discriminating AIH types, AIH-1 (primarily affecting adults), and AIH-2 (primarily affecting children). However, the significance of autoantibodies in determining prognosis, or their role in disease pathogenesis, is debated.

The pathogenesis of AIH is believed to result from a combination of genetic, immunologic, and environmental factors. Regarding genetic predisposition, genome-wide association studies (GWAS) of patients from Europe and North America have shown a significant association between AIH-1 and HLA alleles DRB1*0301 and DRB1*0401^7^. Several studies have further implicated a break-down in self-tolerance as a core immunologic mechanism of disease^8,9^. Numerous environmental triggers have also been associated with AIH, such as medications and viruses, for example minocycline, nitrofurantoin, hepatitis viruses, and human herpes viruses^10,11^. However, a driving, central autoantigen in AIH-1 has not been identified, and the proposed contribution of molecular mimicry as a pathogenic mechanism remains controversial.

While autoantibodies play a central role in AIH diagnosis clinically, further analysis of the pathogenic role of B cells and antibodies would help advance our understanding of AIH, and has been cited as a core goal of the AIH research agenda by professional societies^12^. Phage display immunoprecipitation sequencing (PhIP-seq) is a platform developed to perform an unbiased, high-throughput assessment of antibodies across a broad array of autoimmune conditions^13–21^. We applied PhIP-seq to study serum or plasma from 115 AIH patients obtained from multicenter, international collaborative cohort of patients, and compared these results to a robust series of 298 control serum or plasma samples from patients with metabolic associated steatotic liver disease (MASH), primary biliary cholangitis (PBC), or healthy controls to further define novel disease and tissue specific autoantibody targets to better inform our understanding of AIH pathogenesis.

## Results

### Autoantibody testing was performed on broad, multicenter, international cohorts with robust controls

As part of a multicenter, international collaboration, specimens of serum or plasma from patients with AIH (*n* = 115), MASLD (*n* = 178), or PBC (*n* = 26) and Systemic Sclerosis-Interstitial Lung Disease (SSc-ILD, n = 30), were contributed by four well-characterized patient cohorts: Prospective Observational Study to Understand Liver Diseases (POSULD, San Francisco General Hospital, SF, CA, USA), FrAILT (UCSF Parnassus Hospital, SF, CA, USA), the Eppendorf University cohort (Hamburg, Germany) and the UCSF ILD cohort (UCSF Parnassus Hospital, SF, CA, USA). De-identified healthy control samples were obtained from two sources: the New York Blood Center or purchased through SeraCare (K2EDTA human plasma). Clinical characteristics are summarized in Table 1.

**Table 1.** Patient Characteristics.

| **Table 1a.** Characteristics of the study population assayed by PhIP-seq |  |  |  |  |  |
| --- | --- | --- | --- | --- | --- |
| **Diagnosis** | **n** | **Mean Age** | **Female** | **Caucasian** | **Latinex** |
| AIH | 115 | 58 | 70% | 96% | 14% |
| MASLD | 178 | 60 | 60% | 82% | 67% |
| PBC | 26 | 58% | 94% | 84% | 21% |
| Healthy Control | 94 | -- | -- | -- | -- |

| **Table 1b.** Characteristics of AIH in the patients assayed by PhIP-seq<br>(*n*=115) |  |  |  |  |  |  |  |
| --- | --- | --- | --- | --- | --- | --- | --- |
|  | **F3/4**<br>**fibrosis** | **On**<br>**steroids** | **ANA +** | **ASMA +** | **Mean**<br>**ALT** | **Mean**<br>**AST** | **Mean**<br>**IgG** |
| AIH<br>Patients | 60% | 18% | 88% | 83% | 79 | -- | 1520 |

### A predictive signature for AIH disease status is driven by both novel and previously recognized antibody targets in AIH

A schematic overview of PhIP-seq methodology is summarized in Figure 1a, as well a work-flow for the customized bioinformatic approach to analysis of this data (Fig 1b). Applying these methods, we performed logistic regression with an 85%-15% train-test split ratio with 100 random iterations of cross validation (Fig 1c) using the peptides that were enriched using patient immunoglobin. The resulting model was able to classify a diagnosis of AIH (versus healthy controls) on the basis of PhIP-seq autoreactivity against all peptides (731,724 peptides; median AUC = 0.81). These results were significant, given that serologic diagnosis of AIH is quite heterogenous, including patients who may present without positive autoantibodies at the time of clinical assessment^22^. However, when limiting the logistic regression model to a smaller number of features using only the top 1,000 weighted peptides, predictive power was degraded (median AUC = 0.62), suggesting the presence of substantial heterogeneity in the autoreactivity profiles of individual AIH patients. This suggests that polyreactivity may be a defining feature of autoimmune hepatitis, as previously reported^23^.

**Figure 1.**
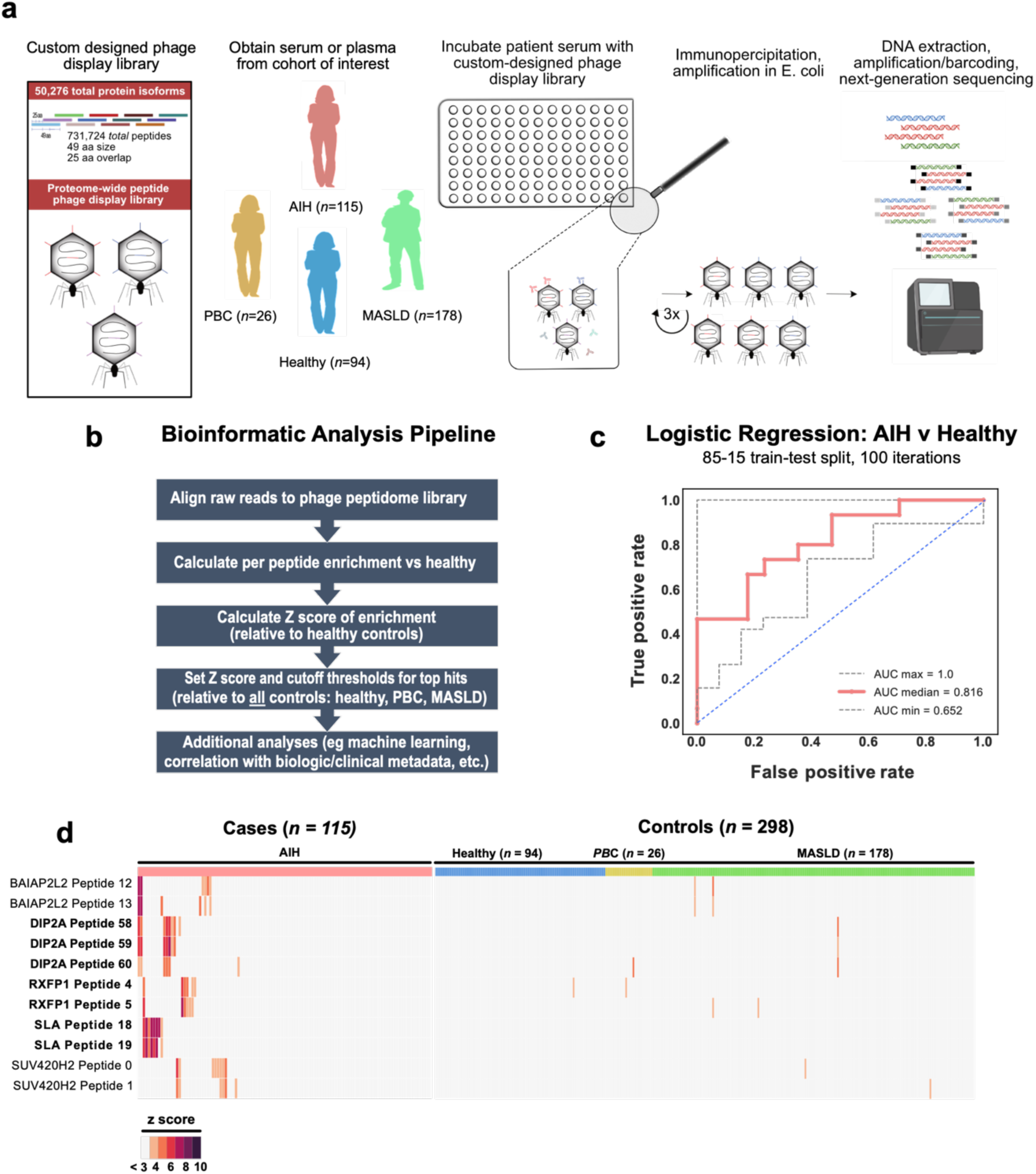
Phage immunoprecipitation sequencing enables AIH disease prediction by logistic regression. (a) Phage display library design, and an overview of methods to apply PhIP-seq to evaluate the autoimmune hepatitis and control cohort. (b) Summary of the customized bioinformatic analysis pipeline to applied to analyze next-generation sequencing output of PhIP-seq data. (c) Receiver operating characteristic (ROC) curve analysis for prediction of AIH vs healthy control disease status, area under the curve (AUC). (d) Heatmap of highly enriched peptides in AIH where multiple overlapping peptides were immunoprecipitated by PhIP-seq (peptide name, left axis). The top legend indicates whether samples correspond to case patients (AIH: pink) or controls (healthy controls: blue, PBC: yellow, MASLD: green); boxes are shaded by Z-score of enrichment (legend bottom left) and Z < 3 is shaded gray. Highlighted in on the left axis in bold are the hits with the top mean Z scores.

Because overfitting is a concern for machine learning techniques, it is important to identify informative features of the model and orthogonally validate them biochemically. We focused our analysis on highly-specific and significantly enriched targets, which were selected by setting a Z-score threshold of >3 relative to the mean of control samples, and further requiring that hits could *not* be significantly enriched in more than 1% of *all* controls (MASLD, PBC, and healthy controls, < 3/298 patients) and must be enriched in at least 5% of the AIH patients (> 6/115 patients). This stringent approach identified 50 hits at the peptide level, representing 44 genes (see Supplemental Table 1 for a complete list of peptides and gene hits meeting criteria). Among the top 50 hits, five proteins were represented by multiple peptides (Figure 1d). This set contained the soluble liver antigen (SLA/LP) protein, a well-characterized AIH autoantigen. Of note, the enriched peptides correspond to the region previously reported to be essential for antibody reactivity to SLA/LP^6^. Four additional proteins are represented in this set, including the relaxin family peptide receptor 1 (RXFP1), the disco-interacting protein 2 homolog A (DIP2A), the lipid binding protein BAIAP2L2, and the methyltransferase SUV420H2 (KMT5C). For additional investigation, we selected the top three proteins by Z-score: SLA/LP, DIP2A, and RXFP1.

The peptides used as bait in the PhIP-seq assay are 49 amino acids in length. To more precisely map the antibody epitopes, a second PhASER (<u>Ph</u>age-<u>A</u>ssisted <u>S</u>canning <u>E</u>pitope <u>R</u>ecovery) PhIP-seq library was created to probe the determinants of antibody binding within selected peptides. To identify the downstream boundary of the SLA/LP epitope, the library included sequential stop-codon substitutions spanning the critical region of reactivity (PhIP-seq SLA/LP peptide 18), employing a similar approach used in our previous investigation of the anti-Hu epitope^18^. Antibody reactivity to the SLA/LP peptide sequence became detectable when the peptide extended through amino acid 414, defining the right-most boundary of the epitope (Figure 2a). This result confirmed the previously reported minimal antigen, and further refined it to a 25 amino stretch (389-414).

**Figure 2.**
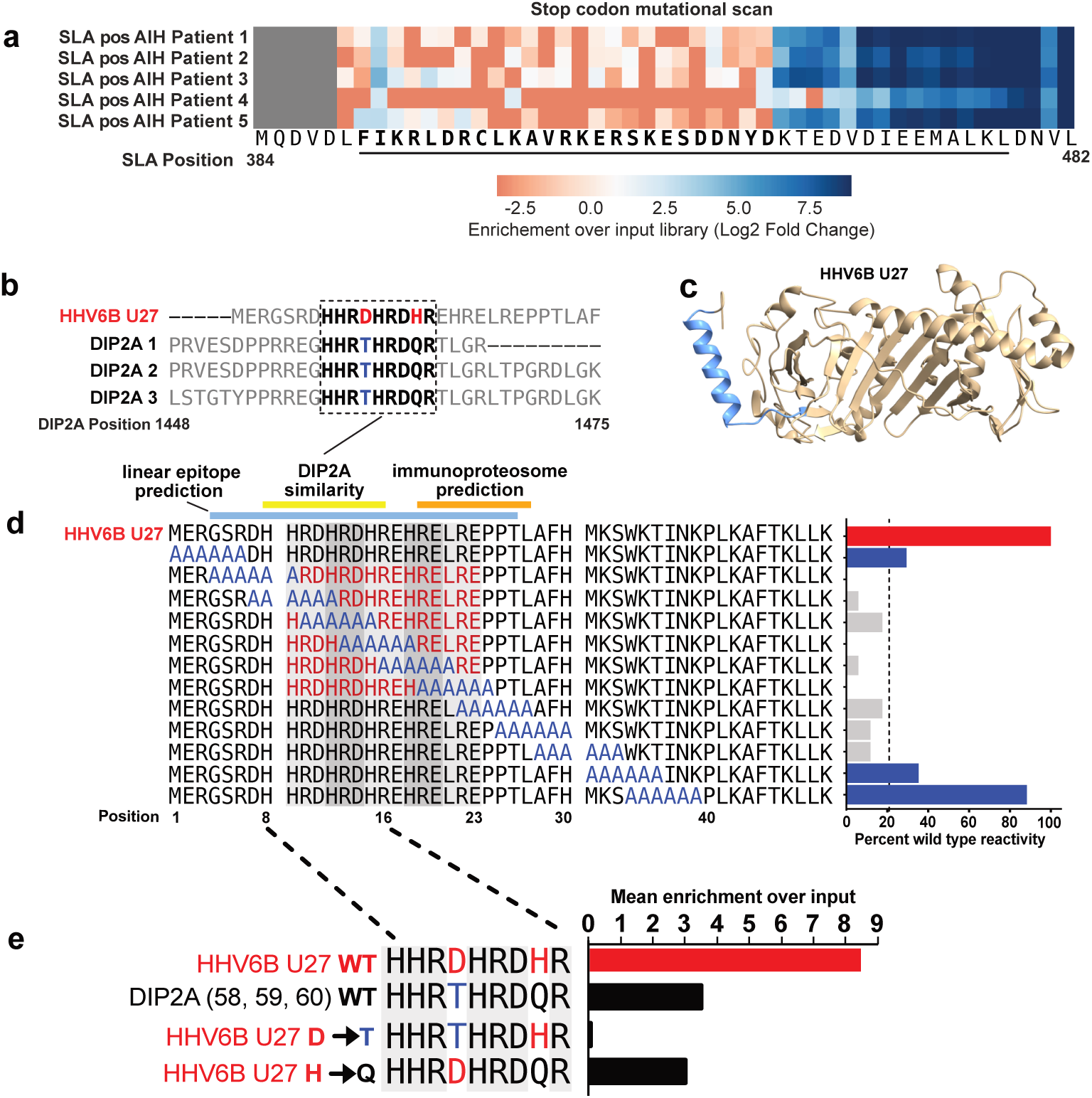
<u>Ph</u>age-<u>A</u>ssisted <u>S</u>canning <u>E</u>pitope <u>R</u>ecovery (PhASER) yields fine mapping of the SLA/LP epitope, and uncovers reactivity to a distinct region of the HHV6 U27 protein with sequence similarity to DIP2A. (a) Heatmap of SLA/LP positive (pos) AIH patient reactivity (n = 5) to SLA/LP peptide 18 (peptide sequence at the bottom of the heatmap) with stop-codon substitutions at each position, starting at SLA/LP amino acid 389. The sequence of the previously-reported minimal SLA/LP epitope is underlined. Reactivity to SLA/LP became detectable (blue) after the peptide extended through position 414, and was not detected with shorter peptide truncations (orange). (b) BLAST search results demonstrating alignment of DIP2A peptide hits to HHV6 U27 protein, 7/9 amino acids were identical, and among non-identical amino acids from U27 (red), only T (blue in DIP2A) ⊒ D (red in U27) is a non-conservative substation. (c) HelixFold rendering of HHV6 U27 protein, with the *bepipred* predicted linear epitope highlighted in blue. (d) HHV6 Alanine scanning schematic (left) and resulting impact on HHV6 reactivity (right); reactivity is normalized to percent of wild type reactivity (red bar). Alanine mutants with <20% of wild type reactivity (gray bars) indicate U27 amino acids 4 – 30 as the critical region for antibody reactivity whereas mutations outside this region (blue bars) had a lesser impact on reactivity. Locations of the *bepipred* predicted linear epitope (blue), DIP2A similarity (yellow), and immunoproteasome processing (orange) all converge on this region of reactivity. (e) Point mutations of the non-identical residues in HHV6 (red) and DIP2A (blue); when reacted with AIH patient serum (*n* = 2), the wild type HHV6 sequence leads to 8x enrichment over input library, more than twice that of DIP2A wild type sequence. Mutating the D found in the HHV6 wild type sequence (red) to the T found in DIP2A (blue; a non-conservative change) abolished this effect.

While hepatic expression of SLA/LP and RXFP1 proteins is well-recognized^24–26^, liver expression of human DIP2A less clear. However, DIP2A was the only protein among the top hits that enriched for 3 overlapping peptides; these 3 peptides share a region of 19 identical amino acids (Figure 2b). Given that any peptide enrichment may be result of antibodies directed against host proteins, or alternatively, against similar non-host sequences, such as a virus, we performed a protein-level BLAST search using the enriched motif as the query. Of the viruses that infect humans, the top match to the DIP2A region of overlap was the U27 protein of human herpes virus 6 (HHV6; Fig 2b). U27 is a viral protein required for replication, which enhances processivity of the viral DNA polymerase, key to the HHV6 lifecycle^27^. The region with sequence similarity to DIP2A is found in the N-terminus of U27 (Fig 2c) and is identical in both HHV6A and HHV6. Interestingly, the SLA/LP protein has also been previously reported to have 41% homology to a peptide derived from the HHV6 U14 protein^28^.

To identify whether antibodies from DIP2A-positive AIH patients react with U27, a PhASER library was employed to display a 49 amino acid stretch of HHV6 U27, spanning the region of similarity. A series of 12 mutated peptides were designed featuring a moving window of 6 consecutive alanine residues, tiled by 3 amino acid steps (Figure 2d). Robust enrichment was observed for the wild type HHV6 peptide sequence among DIP2A positive patients (Figure 2d), not seen in healthy controls. In contrast, when Alanines spanned positions 4 – 30 this enrichment was ablated. Amino acids 4 – 30 directly encompasses the putative similarity region between HHV6 and U27 (residues 8 – 16), identifying this region as central to antibody binding.

To further investigate the contribution of individual amino acid differences in region of putative cross reactivity between DIP2A and the HHV6 U27 sequence (residues 8-16, a subset of the critical region), we used PhASER to perform deep mutational scanning to identify precise determinants of antibody binding, encoding all possible single point mutants at each position of this region (Supplemental Figure 1). This analysis highlighted the importance of the acidic residues. Focusing this approach among the point mutants, the two non-identical positions were mutated from the HHV6 sequence to the DIP2A sequence (Figure 2e). As expected, the semi-conservative change from histidine to glutamine had only a minor impact on immunoprecipitation enrichment with patient sera, whereas the non-conservative change from aspartate to threonine nearly abolished enrichment. This is consistent with the notion that the true target epitope of these antibodies in AIH patients derives from HHV6. Furthermore, no healthy control patients reached our significance threshold for reactivity against DIP2A (z > 3, Supplemental Figure 2a).

HHV6 establishes latency, re-activation has been associated with hepatitis^29^, and HHV-6 infection has been previously associated with the onset of AIH^30^. The critical region of HHV-6 U27 highlighted by the PhASER library has several notable features. First, residues 9-23 are composed of repeating HR[D/E] triplets. Second, *bepipred 2.0* prediction (via IEDB.org) of linear epitopes for U27 surfaced residues 4-26 as a top-ranked candidate (pink bar, Fig 2d). Third, prediction of immunoproteosome processed peptides (for MHC-I presentation) (via IEDB.org) identified residues 19-27 as the top scoring peptide in U27. Given that U27 is an intracellular protein, convergence on both B-cell and T-cell epitopes within the critical region is notable.

### Orthogonal validation of RXFP1 and correlation with patient metadata

Relaxin-2 signaling through RXFP1 on the surface of activated hepatic stellate cells has been shown to decrease their fibrogenic potential^31^, and relaxin-2 has been used as an anti-fibrotic agent in clinical trials of patients with alcohol-associated liver disease^32^. Autoantibody-mediated blockade of RXFP1 signaling, which is anti-fibrotic, has the potential to promote fibrogenesis. Among the 9 AIH patients positive for antibodies against RXFP1 by PhIP-seq, 8 patients (88%) had evidence of advanced fibrosis, F3 or greater.

To validate the RXFP1 finding, we employed a split luciferase binding assay (SLBA), as recently reported^19^. Briefly, *in vitro* transcription and translation was used to generate the primary RXFP1 peptide identified by PhIP-seq with the addition of a HiBiT tag, which when complexed with the LgBiT protein, generates luminescence (Promega system). Immunoprecipitation of tagged peptides was performed with a subset of AIH patients, for which sufficient volumes or serum or plasma were available, in addition to control sera. An antibody targeting the HiBiT protein tag (Promega) was used as a positive control, and negative controls were performed with buffer in the absence of patient serum. A sample was considered positive if the signal in the assay exceeded a cutoff value of the mean plus 3 standard deviations from the mean of all control signal (dotted line, Fig 3a). This assay demonstrated that the same 9 patients positive by PhIP-seq were also positive by SLBA, and none of the liver disease controls or healthy controls were positive (Fig 3a). An additional control of another fibrotic autoimmune disease was included, systemic sclerosis (SSc), as relaxin-2 knockout mice develop systemic fibrosis similar to human SSc and relaxin-based therapy has been pursued in clinical trials of SSc as a therapeutic. We found evidence of only a single positive SSc patient for anti-RXFP1 reactivity among the 30 patients assayed (Fig 3, green), further suggesting anti-RXFP1 antibodies are associated specifically with AIH as opposed to other autoimmune conditions. Further investigation of the 9 RXFP1(+) patients (Z-score > 2.5) had some evidence of disease activity (defined by AST or ALT greater than 2x the upper limit of normal OR an elevated IgG level), and 8/9 patients (88%) had evidence of advanced fibrosis, F3 or greater.

**Figure 3.**
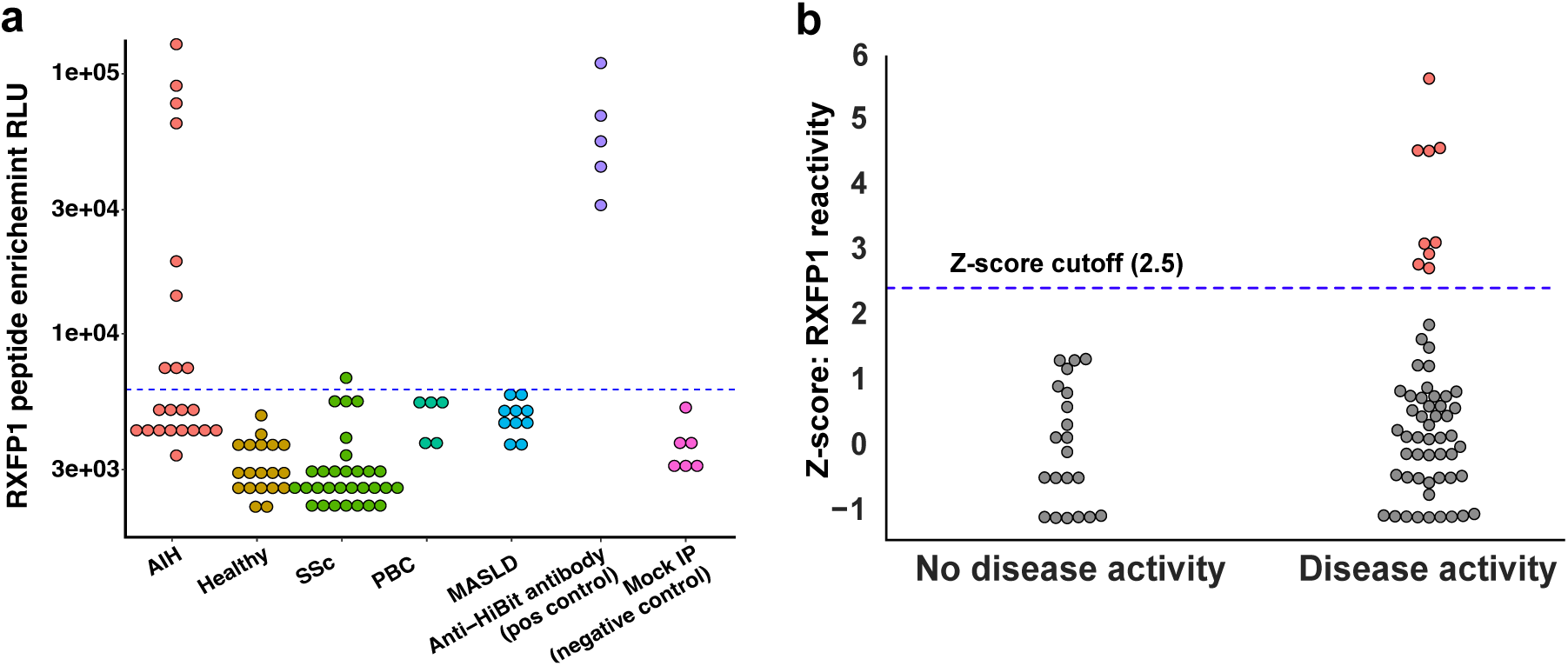
Orthogonal validation of RXFP1 rectivity, and correlation with AIH disease activity. (a) SLBA validation of anti-RXFP1 peptide reactivity in various patients groups (x-axis), as measured by enrichment of relative light units (RLU, y-axis); the cutoff for positivity was set at the mean + 3 standard deviations of all controls (blue dashed line). (b) PhIP-seq data plotting Z-score of anti-RXFP1 peptide reactivity among AIH patients (y-axis); AIH patients were separated into groups of active vs inactive AIH (x-axis); patients considered as positivity reactive against anti-RXFP1 had a Z-score of enrichment >2.5 (denoted by blue dashed line, highlighted in pink).

### Serum from AIH patients with anti-RXFP1 activity inhibits relaxin-2 signaling through RXFP1 in an IgG-dependent manner

The functional implications of RXFP1 positivity were explored further to investigate the possibility that the autoantibody itself is pathogenic in AIH. The proposed structure of RXFP1 is diagrammed in Fig 4a using UCSF ChimeraX^33^, with annotation of various domains. The extracellular portion of RXFP1 (light gray, Fig 4a) is composed primarily of a large, leucine-rich repeat (LRR) motif, present among a larger family of LRR containing G protein-coupled receptors (LGRs)^34^. This LRR repeat region is the site of relaxin-2 ligand binding. Further highlighted in red is the RXFP1 peptide target of antibodies in patient serum, as identified in PhIP-seq and SLBA assays. At the core of this peptide is an LRRNT motif, an N-terminal capping region required to maintain stability of the leucine-rich repeat (LRR) region. The RXFP1 peptide is at the end of a linker region connecting the LRRNT to a low-density lipoprotein domain (LDLa domain; diagrammed in red, Fig 4a inset panel). This linker has been demonstrated to be required for RXFP1 receptor activation^35^. Epitope mapping by PhASER further narrowed the epitope to a region of 7 critical amino acids (Supplemental Figure 3). Based on the peptide’s position within RXFP1, we hypothesized that antibodies targeting this epitope would be able to functionally interfere with RXFP1 signaling. We tested this hypothesis using the cAMP-Glo signaling assay (Promega) in which ligand binding to its cognate G protein-coupled receptor (GPCR) leads to the production of cAMP, which can be measured by luminescence as a sensitive read-out of ligand binding to the GPCR. We performed this assay in THP-1 cells, a human monocytic cell line, as cAMP production in response to relaxin-2 binding of RXFP1 has been well characterized in this system^36^.

**Figure 4.**
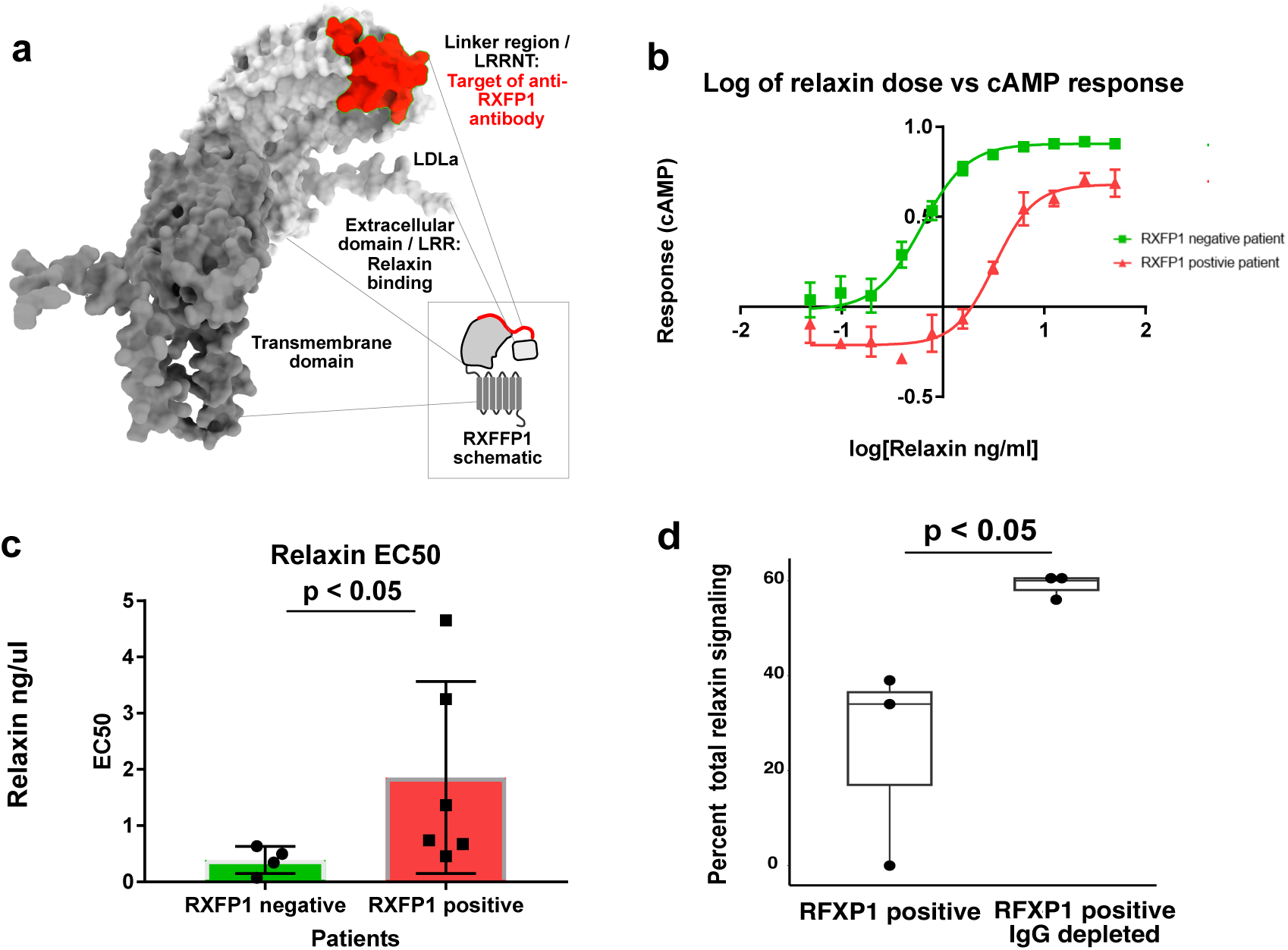
Serum from AIH patients with anti-RXFP1 activity inhibits relaxin-2 signaling through RXFP1 in an IgG-dependent manner. (a) Putative structure of RXFP1, as depicted using ChimeraX; the region corresponding to the RXFP1 peptide identified by PhIP-seq is highlighted in red, along with annotation of functional domains (for schematic representation, see panel inset). (b) Assay of relaxin-2-induced induction of cAMP by RXFP1, in TH1 cells pre-incubated with [1:100] dilution of patient serum negative (green) or positive (red) for RXFP1 antibodies; relaxin concentration (x-axis), cAMP response reported as a percent of untreated control signal, y-axis. (c) Measurement of relaxin-2 EC50 in ng/µl (y-axis) for patient serum negative (green) or positive (red) for RXFP1 antibodies. (d) Depletion of IgG using protein A-G beads (x-axis, right) or mock-depleted serum (x-axis, left) was performed prior to incubating THP-1 cells with patient serum at [1:250]; resultant impact on relaxin-2 signal expressed as a percent of untreated signal (y-axis).

A dose response curve was first created using THP-1 cells incubated with serum (2 hours) from an anti-RXFP1-positive and an anti-RXFP1-negative patient. Treatment with anti-RXFP1-positive serum resulted in a significant shift in the EC50 (5-fold) (Fig 4b). Repeating this assay on 5 additional patients and 4 additional controls revealed a significant difference over a range of values (Fig 4c). AG-bead based depletion of IgG prior to pre-incubation with anti-RXFP1 positive serum abrogated this effect (Fig 4d), consistent with the notion that the inhibitory component in patient serum is indeed IgG immunoglobins.

## Discussion

AIH has been rising in incidence and prevalance^37^, and despite this trend, the etiopathogenesis of the disease remains incompletely understood. Environmental triggers have been repeatedly implicated in AIH pathogenesis, including infections^38^. There are also several experimental models of the AIH that require an infectious trigger to overcome the immune tolerant environment in the liver^39^. However, despite this long-standing association between AIH and pathogens such as HHV6^6^, direct molecular evidence is lacking. Furthermore, despite a growing list of biological processes associated with AIH^40^, advances in disease therapeutics remain elusive^40^. Novel approaches are required to accelerate our understanding of AIH pathogenesis.

Here, we used PhIP-seq to uncover novel autoantibody targets in AIH patients, by leveraging a large cohort of patients from an international, multi-center collaboration to identify new aspects of AIH pathophysiology. One unique aspect of this cohort is the inclusion of many non-AIH liver disease controls including 178 patients with MASLD. MASLD is a disease that can coexist with AIH, and can be difficult to differentiate from AIH on the basis of serology given the high rates of co-occurring autoantibodies, such as ANA and anti-smooth muscle antibodies, reported to be positive in 20-30% of MASLD patients^4,41^. Our study identified several antibodies highly-specific Analysis of the aggregated PhIP-seq data using machine learning, comparing AIH to healthy controls by logistic regression, yielded an average AUC of 0.81. These data support the notion that shared autoreactivities exist within subsets of AIH patients. While these data suggest predictive potential, a larger, extensively characterized multicenter validation cohort would be required to further validate the approach as a diagnostic.

To further leverage the value of PhIP-seq performed on patients from a multi-center international cohort of AIH patients and controls, we focused on the identification of disease- and tissue-specific antibodies. SLA/LP, a known autoantigen in AIH, was identified as the most specific target using this approach, and thus served as an internal control. RXFP1 has not been previously described in AIH and was enriched by a subset of patient sera, but not in healthy controls. Furthermore, serum from anti-RXFP1 positive patients inhibits relaxin-2 signaling via RXFP1, suggesting the antibodies directed against this protein may play a direct functional role. Specifically, our results suggest that anti-RXFP1 antibodies have potential to diminish the anti-fibrotic properties of relaxin-2. Additional studies will be required to better understand the role of these antibodies and whether they may enable risk stratification of AIH patients for fibrosis progression.

A third autoreactivity was initially ascribed to the protein DIP2A, which is not known to have a specific role in the liver. Investigation revealed that the autoreactive region of DIP2A shares significant sequence similarity to the HHV-6 processivity factor U27. Through mutagenesis, the critical region was further refined to encompass a 22 amino acid segment near the N-terminus of U27. Individual point mutations further support the notion that the observed autoreactivity to DIP2A was due to antibodies directed against HHV-6 U27. This segment harbors several triplet repeats, and it also contains predicted linear epitopes as well as a predicted immunoproteasome fragment. To our knowledge, this is the first evidence that AIH patients positive for highly-specific autoantibodies also react with increased reactivity to a highly-similar region of viral protein. While the seroprevalence of HHV-6 in the adult population can exceed 90%^42^, immunoreactivity to this segment of U27 is likely to be uncommon, as no healthy controls yielded enrichment of this fragment. This selective immune response may be attributable to HLA-restricted autoreactive T cells with cross reactive potential, similar to what was recently described for SARS-Cov-2 and MIS-C^43^. While beyond the scope of this initial study, further investigation, including T cell epitope profiling using patient PBMCs, could yield additional mechanistic insights.

This study has several limitations. Analysis by logistic regression suggests that a multitude of features are required to classify AIH samples from healthy subjects, which implies that shared autoreactive targets are only common in smaller subsets within the cohort. Thus, the ability to positively identify additional shared autoreactivities is limited by the overall cohort size. Indeed, the combined contributions of SLA/LP, RXFP1, and HHV-6 U27 are positive in less than 1/3 of the AIH samples analyzed here. In addition, the PhIP-seq technique employed here largely captures linear epitopes. Conformational epitopes, post translationally modified epitopes, and other non-linear configurations would likely be undetected in these assays. Ongoing studies, including additional methods of autoantibody repertoire screenings, investigation of T cell epitopes, and cloning of antigen specific B cells from patient PBMCs would all contribute to a better understanding of AIH and may also help to identify therapeutic targets.

## Data Availability

All PhIP-seq data are available for download at Dryad.

## Acknowledgements

Thanks to D. Montgomery Bissell, Emily D. Crawford, Michael Wilson for their thoughtful contributions to orchestrating collaborations and getting this project off the ground.

## Patients and Methods

### Patient cohorts

As part of a multicenter, international collaboration, specimens of serum or plasma from patients with AIH (*n* = 115), MASLD (*n* = 178), or PBC (*n* = 26) and SSc-ILD (n = 30), were contributed by four well-characterized patient cohorts: Prospective Observational Study to Understand Liver Diseases (POSULD, San Francisco General Hospital, SF, CA, USA), FrAILT (UCSF Parnassus Hospital, SF, CA, USA), the Eppendorf University cohort (Hamburg, Germany) and the UCSF ILD cohort (UCSF Parnassus Hospital, SF, CA, USA). All patients provided informed consent in accord with institutional policies. Clinical data were obtained from existing databases and the medical record, including demographic information and disease activity, fibrosis stage, and medication regimen near the time of specimen collection. Patients with more than one diagnosis (eg overlap syndromes, AIH and MASLD, etc.) were excluded from the current analysis. Coded specimens were analyzed in a deidentified fashion. Healthy control samples were obtained as de-identified samples from two sources: the first was from the New York Blood Center, as part of retention tubes collected at the time of blood donation from volunteer donors who provided their informed consent for their samples to be used for research. The second source was patient plasma from donors obtained from FDA-licensed blood collection facilities, purchased through SeraCare (K2EDTA human plasma).

### Phage immunopercipitaion sequencing (PhIP-seq)

PhIP-seq was performed as previously reported^15,16,44^ and PhIP-seq protocols described in detail are available at protocols.io, with a multichannel-based scaled protocol (https://www.protocols.io/view/scaled-moderate-throughput-multichannel-phip-proto-8epv5zp6dv1b/v1) and a stand-alone guide to library preparation (https://www.protocols.io/view/phage-display-library-prep-method-rm7vz3945gx1/v1).

Briefly, blood from individuals with Type 1 AIH and controls was analyzed using PhIP-seq. Phage were cloned to express >700,000 overlapping peptides spanning the human proteome. The following text adapted from Zamecnik *et al*^13^: 96-well, 2mL deep well polypropylene plates were incubated with a blocking buffer (3% BSA in TBST) overnight at 4°C to prevent nonspecific binding. Blocking buffer was then replaced with 500 µL of freshly grown phage library and 1 µL of human sera diluted 1:1 in storage buffer (PBS with 0.04% NaN3, 40% Glycerol, 40mM HEPES). To facilitate antibody-phage binding, the deep well plates with library and sample were incubated overnight at 4°C on a rocker platform. 10 µL of each of Pierce Protein A and G Beads (ThermoFisher Scientific, 10002D & 10004D) slurry were aliquoted per reaction and washed 3 times in TNP-40 (140mM NaCl, 10mM Tris-HCL, 0.1% NP40). After the final wash, beads were resuspended in TNP-40 in half the original slurry volume (20uL) and added to the phage-patient antibody mixture and incubated on the rocker at 4°C for 1 hour. Beads were then washed in RIPA buffer, and then the immunoprecipitated solution was resuspended in 150 µL of LB-Carb and then added to 0.5mL of log-phase BL5403 E. coli for amplification (OD600 = 0.4-0.6) until lysis was complete (approximately 2h) on an 800 rpm shaker. After amplification, sterile 5M NaCl was added to lysed E. coli to a final concentration of 0.5M NaCl to ensure complete lysis. The lysed solution was spun at 3220 rcf for 20 minutes and the top 500 µL was filtered to remove remaining cell debris. Filtered solution was transferred to a new pre-blocked deep-well plate where patient sera was added and subjected to another round of immunoprecipitation and amplification, and 3 total rounds of immunoprecipitation were completed. The final lysate was spun at 3220 xg for 30 minutes, with supernatant then filtered and stored at 4°C for subsequent NGS library prep. Phage DNA from each sample was barcoded and amplified (Phusion PCR) and then underwent Next-Generation Sequencing on an Illumina NovaSeq Instrument (Illumina, San Diego, CA).

### Split Luciferase Binding Assay (SLBA)

This assay was performed as recently reported^19^; and a detailed SLBA protocol is available on protocols.io at dx.doi.org/10.17504/protocols.io.4r3l27b9pg1y/v1.

Briefly, the target peptide of relaxin family peptide was identified by PhIP-seq, with the following peptide sequence: VGSVPVQCLCQGLELDCDETNLRAVPSVSSNVTAMSLQWNLIRKLPPDC. The following text was adapted from Rackaityte *et al*.^19^: The nucleic acid sequence of this construct was inserted into a split luciferase construct containing a terminal HiBiT tag and synthesized (IDT) as DNA oligomers. Constructs were amplified by PCR using 5’-AAGCAGAGCTCGTTTAGTGAACCGTCAGA-3’ and 5’-GGCCGGCCGTTTAAACGCTGATCTT-3’ primer pair. Unpurified PCR product was used as input to rabbit reticulocyte transcription translation system (Promega) and Nano-Glo HiBit Lytic Detection System (Promega Cat No. N3040) was used to measure relative luciferase units (RLU) of translated peptides in a luminometer. Peptides were normalized to 5e6 RLU input, incubated overnight with patient sera, and immunoprecipitated with protein A and protein G sepharose beads (Millipore Sigma). After thoroughly washing beads with SLBA buffer (0.15M NaCl, 0.02M Tris-HCl pH7.4, 1% w/v sodium azide, 1% w/v bovine serum albumin, and 0.15% v/v Tween-20), luminescence remaining on beads was measured using Nano-Glo HiBit Lytic Detection System (Promega Cat No. N3040) in a luminometer. Anti-HiBiT antibody (Promega) was used as a positive control for each peptide. A patient was considered positive by SLBA if the RLU exceeded the mean of all controls + 3 standard deviations.

### Relaxin-2 signaling/cAMP-Glo assay/IgG depletion

Recombinant human relaxin H2 was purchased from R&D systems (catalog # 6586-RN-025/CF) and resuspended in 1x sterile PBS with 1% BSA at a concentration of 100 µl/ml. In order to block cyclic nucleotide phosphodiesterases during the cAMP-Glo assay, serial dilutions of relaxin were made up in induction buffer composed of 1x PBS with 500 µM 3-isobutyl-1-methylxanthine (IBMX, Sigma Aldrich), and 500 µM Ro 20-1724 (Cayman Chemical). Relaxin concentrations ranged from 0.0488 – 50 ng/ul of ligand, and the 12^th^ dilution was “untreated” control, of just induction buffer. Dilutions were made in sterile 96 well plates in order to apply to THP1 cells to study signaling. THP1 cells were obtained via ATCC, and seeded at a density of 1×10^6^ cells/well of a 96-well plate. Prior to the addition of relaxin-2, THP1 cells were pre-incubated with patient serum from RXFP1 positive patients or RXPF1 negative patients at a dilution of [1:100] in RPMI with 10% FBS/1% PSG for 2 hours in a 37 °C incubator. Following this pre-incubation, relaxin was added at each of the 11 pre-diluted concentrations to pre-incubated THP-1 cells, and 96-well plates were returned to the 37 °C incubator for two hours. All reactions were performed in triplicate. Following this incubation, cells were assayed for cAMP production using the cAMP-Glo assay (Promega catalog # V1502) was performed per the manufacturer’s instructions, with the following modifications. Lysis of cells was performed using 20 µl of cAMP-Glo lysis buffer for 30 minutes in standard tissue culture plates, and then transferred to opaque white 96-well plates (Nunc) for the remainder of the assay in order to facilitate plate reading of relative light units (RLU) in a luminometer (Promega). The remainder was as per manufacturer’s instructions. Results were then normalized to the fraction of untreated RLU, and EC50 values were calculated using GraphPad Prism software. For depletion experiments, prior to cAMP-Glo assay, as described above, sera was pre-incubated for 2 hours at room temperature with Pierce Protein A and G Beads (Pierce) at a ratio of 20 µl of A/20 µl G to 1 µl of serum, with gentle rocking. Samples were ultimately pre-incubated with THP1 cells at a dilution of [1:250] prior to addition of relaxin-2.

### Bioinformatic and statistical analysis

Raw sequencing reads were aligned to the input peptide library using RAPsearch2 (protein alignment), as previously described^16^. Aligned reads were controlled for varying read depth by normalizing reads per 100,000 (RPK). Normalized PhIPseq read counts were further analyzed using Python. Code to perform phage data analysis can be found in the PhagePy Package (https://github.com/h-s-miller/phagepy).

Briefly, fold-change for each peptide was generated relative to mean RPK of the controls, and the Z-score was calculated from the background distribution. An peptide was considered a hit if enrichment was >3 standard deviations from the mean of healthy controls and present in at least 6 AIH patients (5% sensitivity) and not present in more than 1% of control patients (99% specificity). Logistic regression was performed using the Scikit-learn package in Python^45^, using the liblinear solver and L1 regularization. The model was evaluated with and 85-15 train-test-split ratio, and performed with 100 iterations of cross validation. Molecular graphics and analyses performed with UCSF ChimeraX (https://www.cgl.ucsf.edu/chimerax/), developed by the Resource for Biocomputing, Visualization, and Informatics at the University of California, San Francisco, with support from National Institutes of Health R01-GM129325 and the Office of Cyber Infrastructure and Computational Biology, National Institute of Allergy and Infectious Diseases. HLA Class II binding predictions were performed using the Immune Epitope Database (IEDB, www.iedb.org)^46^.

## Supplemental Figures

**Supplemental Table 1.** Top 50 peptide hits identified by PhIP-seq.

|  | GI Number | gene | pep peptide short | sensitivity | specificity<br>(relative to all controls) |
| --- | --- | --- | --- | --- | --- |
| 1 | gi\|768020101 | ADARB1 | ADARB1 peptide 22 | 5.22% | 99.33% |
| 2 | gi\|170650694 | AGAP2 | AGAP2 peptide 38 | 5.22% | 100.00% |
| 3 | gi\|578838409 | ATRX | ATRX peptide 63 | 5.22% | 99.33% |
| 4 | gi\|768025790 | BAIAP2L2 | BAIAP2L2 peptide 12 | 5.22% | 99.33% |
| 5 | gi\|530420394 | BAIAP2L2 | BAIAP2L2 peptide 13 | 5.22% | 99.33% |
| 6 | gi\|38788274 | BPTF | BPTF peptide 100 | 5.22% | 99.66% |
| 7 | gi\|24431975 | C10orf12 | C10orf12 peptide 44 | 5.22% | 99.33% |
| 8 | gi\|62339432 | C10orf54 | C10orf54 peptide 10 | 5.22% | 99.33% |
| 9 | gi\|767938327 | C5orf60 | C5orf60 peptide 8 | 5.22% | 100.00% |
| 10 | gi\|16554564 | CARD6 | CARD6 peptide 42 | 6.09% | 99.33% |
| 11 | gi\|586798144 | CECR2 | CECR2 peptide 38 | 5.22% | 100.00% |
| 12 | gi\|96975038 | CEP120 | CEP120 peptide 14 | 5.22% | 99.66% |
| 13 | gi\|54112403 | CHD7 | CHD7 peptide 21 | 6.96% | 99.33% |
| 14 | gi\|530420632 | DEPDC5 | DEPDC5 peptide 26 | 5.22% | 99.66% |
| 15 | gi\|768015660 | DIDO1 | DIDO1 peptide 93 | 5.22% | 99.33% |
| 16 | gi\|768020426 | DIP2A | DIP2A peptide 58 | 6.96% | 99.66% |
| 17 | gi\|768020426 | DIP2A | DIP2A peptide 59 | 6.09% | 99.66% |
| 18 | gi\|768020424 | DIP2A | DIP2A peptide 60 | 5.22% | 99.33% |
| 19 | gi\|383387816 | EME2 | EME2 peptide 14 | 5.22% | 99.66% |
| 20 | gi\|530364644 | FCRL6 | FCRL6 peptide 3 | 5.22% | 100.00% |
| 21 | gi\|34452723 | GABRA4 | GABRA4 peptide 16 | 5.22% | 99.33% |
| 22 | gi\|7019419 | GNL2 | GNL2 peptide 18 | 5.22% | 99.33% |
| 23 | gi\|148612838 | KIAA2026 | KIAA2026 peptide 81 | 6.09% | 99.33% |
| 24 | gi\|308199413 | KMT2A | KMT2A peptide 43 | 5.22% | 99.33% |
| 25 | gi\|767975406 | KMT2D | KMT2D peptide 147 | 5.22% | 99.66% |
| 26 | gi\|767929517 | LOC101928621 | LOC101928621 peptide 17 | 5.22% | 99.33% |
| 27 | gi\|768040476 | LOC105373377 | LOC105373377 peptide 6 | 6.09% | 100.00% |
| 28 | gi\|731441417 | MARK3 | MARK3 peptide 16 | 6.09% | 99.66% |
| 29 | gi\|767925635 | MED12L | MED12L peptide 79 | 5.22% | 99.66% |
| 30 | gi\|256017159 | MGA | MGA peptide 99 | 5.22% | 100.00% |
| 31 | gi\|45356151 | NCAPD3 | NCAPD3 peptide 55 | 5.22% | 99.66% |
| 32 | gi\|221139926 | NHSL1 | NHSL1 peptide 62 | 5.22% | 99.66% |
| 33 | gi\|54144631 | PHACTR1 | PHACTR1 peptide 19 | 5.22% | 99.33% |
| 34 | gi\|115430211 | PHACTR4 | PHACTR4 peptide 24 | 5.22% | 99.66% |
| 35 | gi\|530425349 | PTPRS | PTPRS peptide 33 | 5.22% | 99.66% |
| 36 | gi\|530372696 | QRICH1 | QRICH1 peptide 17 | 5.22% | 99.33% |
| 37 | gi\|148613886 | RFX7 | RFX7 peptide 20 | 6.09% | 99.33% |
| 38 | gi\|209915591 | RPS6KC1 | RPS6KC1 peptide 30 | 6.09% | 99.33% |
| 39 | gi\|767932471 | RXFP1 | RXFP1 peptide 4 | 5.22% | 99.33% |
| 40 | gi\|767932469 | RXFP1 | RXFP1 peptide 5 | 5.22% | 99.33% |
| 41 | gi\|767929800 | SEPSECS | SLA/LP peptide 18 | 6.96% | 100.00% |
| 42 | gi\|767929798 | SEPSECS | SLA/LP peptide 19 | 6.09% | 100.00% |
| 43 | gi\|530416300 | SIGLECL1 | SIGLECL1 peptide 6 | 5.22% | 99.66% |
| 44 | gi\|767916325 | SPEG | SPEG peptide 63 | 5.22% | 99.33% |
| 45 | gi\|768011637 | SUV420H2 | SUV420H2 peptide 0 | 6.96% | 99.66% |
| 46 | gi\|768011637 | SUV420H2 | SUV420H2 peptide 1 | 5.22% | 99.66% |
| 47 | gi\|7662078 | TTC37 | TTC37 peptide 57 | 5.22% | 99.66% |
| 48 | gi\|4507779 | UBE2E1 | UBE2E1 peptide 1 | 5.22% | 99.66% |
| 49 | gi\|238550206 | ZBP1 | ZBP1 peptide 16 | 5.22% | 99.33% |
| 50 | gi\|545479119 | ZNF133 | ZNF133 peptide 6 | 5.22% | 99.33% |

**Supplemental Figure 1.**
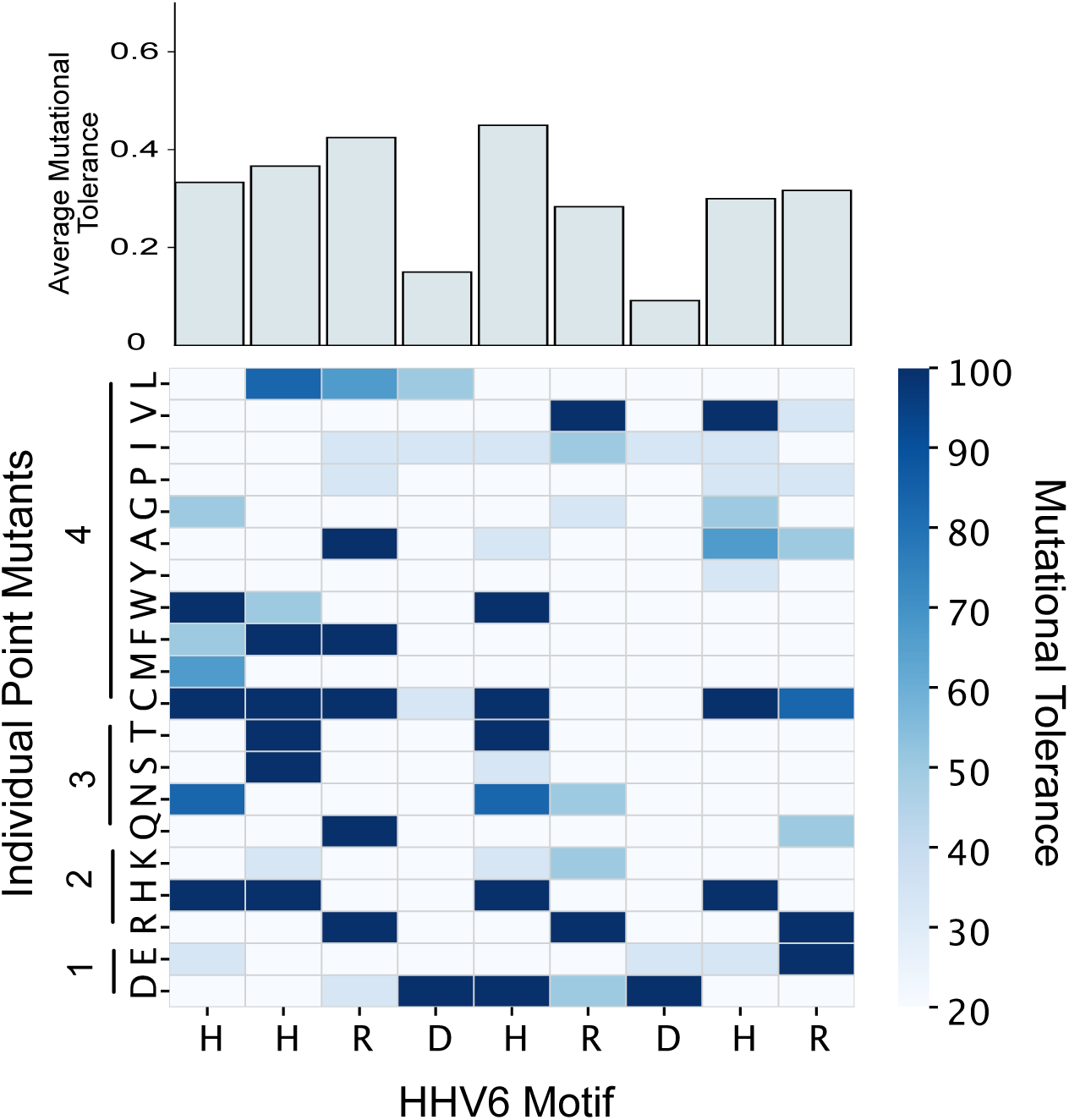
Deep mutational scanning of the cross-reactive motif between DIP2A and HHV6 U27. The heatmap denotes the impact of point mutation at each position in the motif HHV6 U27 motif, residues 8-16 (x-axis) relative to each point mutant (y-axis), which are grouped by properties of their side chains (1 – acidic/negative charge, 2 – basic/positive charge, 3 – polar, uncharged and 4 – hydrophobic). Data are expressed as a percent of the wild-type U27 reactivity (mutational tolerance, legend at right). Average mutational tolerance at each position of the motif is summarized at the top of the heatmap.

**Supplemental Figure 2.**
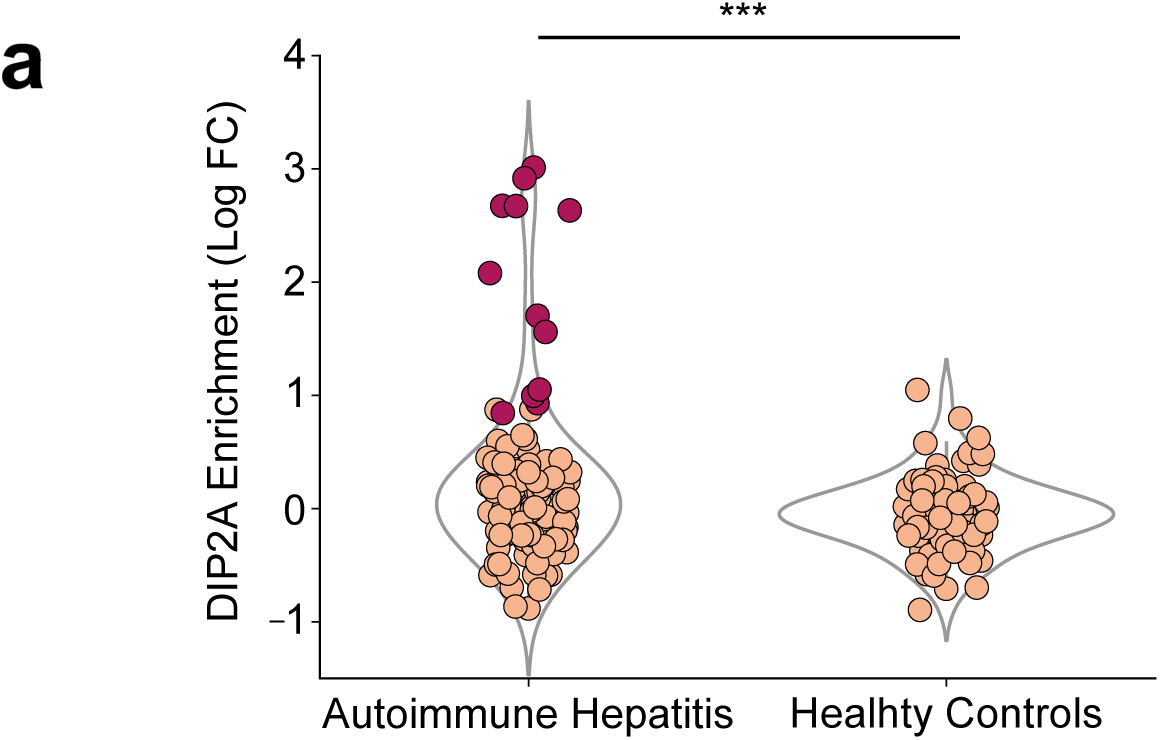
Enrichment in DIP2A by PhIP-seq. Enrichment in AIH vs healthy Controls (x-axis), measured by Log fold change of DIP2A relative to the mean of DIP2A enrichment in healthy control samples (y-axis), where patients with a z-score of >3 relative to the mean of healthy controls in colored in dark red (***p < 0.01).

**Supplemental Figure 3.**
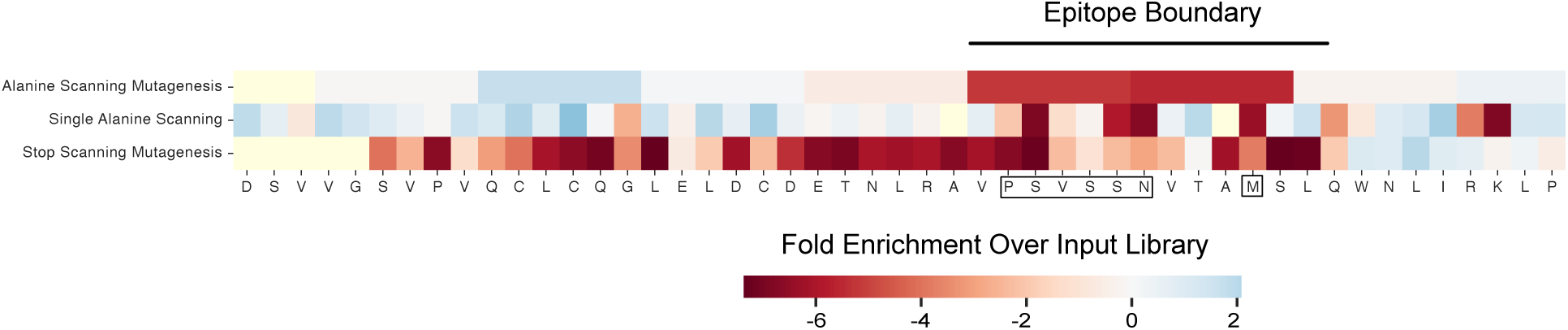
PhASER epitope mapping of RXFP1. The RXFP1 peptide is noted at the bottom, with the identified epitope outlined (black box), identified by areas where mutagenesis caused a loss or reactivity (red) as measured by fold enrichment over input library using various approaches denoted on the left axis (single alanine scanning and 6-mer blocks of alanine scanning) as well as stop scanning mutagenesis, used to define the epitope boundary (line above the figure).

## References

1. Lamba M, Ngu JH, Stedman CAM. Trends in Incidence of Autoimmune Liver Diseases and Increasing Incidence of Autoimmune Hepatitis. Clinical Gastroenterology and Hepatology. Elsevier; 2021 Mar 1;19(3):573–579.e1. PMID: 32526342

2. Lee B, Holt EW, Wong RJ, Sewell JL, Somsouk M, Khalili M, Maher JJ, Tana MM. Race/ethnicity is an independent risk factor for autoimmune hepatitis among the San Francisco underserved. Autoimmunity. 2018 Aug;51(5):258–264. PMCID: PMC6311708

3. Wen JW, Kohn MA, Wong R, Somsouk M, Khalili M, Maher J, Tana MM. Hospitalizations for Autoimmune Hepatitis Disproportionately Affect Black and Latino Americans. Am J Gastroenterol. 2018 Feb;113(2):243–253. PMCID: PMC6522224

4. Kanzler S, Weidemann C, Gerken G, Löhr HF, Galle PR, Büschenfelde KHM zum, Lohse AW. Clinical significance of autoantibodies to soluble liver antigen in autoimmune hepatitis. Journal of Hepatology. 1999 Oct 1;31(4):635–640.

5. Manns M, Kyriatsoulis A, Gerken G, Staritz M, Meyer KH, Büschenfelde Z. CHARACTERISATION OF A NEW SUBGROUP OF AUTOIMMUNE CHRONIC ACTIVE HEPATITIS BY AUTOANTIBODIES AGAINST A SOLUBLE LIVER ANTIGEN. The Lancet. 1987 Feb 7;329(8528):292–294.

6. Wies I, Brunner S, Henninger J, Herkel J, Kanzler S, zum Büschenfelde KHM, Lohse AW. Identification of target antigen for SLA/LP autoantibodies in autoimmune hepatitis. The Lancet. 2000 Apr 29;355(9214):1510–1515.

7. de Boer YS, van Gerven NMF, Zwiers A, Verwer BJ, van Hoek B, van Erpecum KJ, Beuers U, van Buuren HR, Drenth JPH, den Ouden JW, Verdonk RC, Koek GH, Brouwer JT, Guichelaar MMJ, Vrolijk JM, Kraal G, Mulder CJJ, van Nieuwkerk CMJ, Fischer J, Berg T, Stickel F, Sarrazin C, Schramm C, Lohse AW, Weiler-Normann C, Lerch MM, Nauck M, Völzke H, Homuth G, Bloemena E, Verspaget HW, Kumar V, Zhernakova A, Wijmenga C, Franke L, Bouma G, Dutch Autoimmune Hepatitis Study Group, LifeLines Cohort Study, Study of Health in Pomerania. Genome-wide association study identifies variants associated with autoimmune hepatitis type 1. Gastroenterology. 2014 Aug;147(2):443–452.e5. PMID: 24768677

8. Longhi MS, Ma Y, Bogdanos DP, Cheeseman P, Mieli-Vergani G, Vergani D. Impairment of CD4(+)CD25(+) regulatory T-cells in autoimmune liver disease. J Hepatol. 2004 Jul;41(1):31–37. PMID: 15246204

9. Ferri S, Longhi MS, De Molo C, Lalanne C, Muratori P, Granito A, Hussain MJ, Ma Y, Lenzi M, Mieli-Vergani G, Bianchi FB, Vergani D, Muratori L. A multifaceted imbalance of T cells with regulatory function characterizes type 1 autoimmune hepatitis. Hepatology. 2010 Sep;52(3):999–1007. PMID: 20683931

10. Zachou K, Arvaniti P, Lyberopoulou A, Dalekos GN. Impact of genetic and environmental factors on autoimmune hepatitis. J Transl Autoimmun. 2021 Sep 21;4:100125. PMCID: PMC8479787

11. Manns MP, Lohse AW, Vergani D. Autoimmune hepatitis – Update 2015. Journal of Hepatology. Elsevier; 2015 Apr 1;62(1):S100–S111. PMID: 25920079

12. Autoimmune hepatitis: From current knowledge and clinical practice to future research agenda - Sebode - 2018 - Liver International - Wiley Online Library [Internet]. [cited 2023 Jun 11]. Available from: https://onlinelibrary.wiley.com/doi/10.1111/liv.13458

13. Zamecnik CR, Sowa GM, Abdelhak A, Dandekar R, Bair RD, Wade KJ, Bartley CM, Tubati A, Gomez R, Fouassier C, Gerungan C, Alexander J, Wapniarski AE, Loudermilk RP, Eggers EL, Zorn KC, Ananth K, Jabassini N, Mann SA, Ragan NR, Santaniello A, Henry RG, Baranzini SE, Zamvil SS, Bove RM, Guo CY, Gelfand JM, Cuneo R, Büdingen HC von, Oksenberg JR, Cree BA, Hollenbach JA, Green AJ, Hauser SL, Wallin MT, DeRisi JL, Wilson MR. A Predictive Autoantibody Signature in Multiple Sclerosis [Internet]. medRxiv; 2023 [cited 2023 Jun 11]. p. 2023.05.01.23288943. Available from: https://www.medrxiv.org/content/10.1101/2023.05.01.23288943v1

14. Larman HB, Zhao Z, Laserson U, Li MZ, Ciccia A, Gakidis MAM, Church GM, Kesari S, Leproust EM, Solimini NL, Elledge SJ. Autoantigen discovery with a synthetic human peptidome. Nat Biotechnol. 2011 May 22;29(6):535–541. PMCID: PMC4169279

15. Vazquez SE, Mann SA, Bodansky A, Kung AF, Quandt Z, Ferré EM, Landegren N, Eriksson D, Bastard P, Zhang SY, Liu J, Mitchell A, Proekt I, Yu D, Mandel-Brehm C, Wang CY, Miao B, Sowa G, Zorn K, Chan AY, Tagi VM, Shimizu C, Tremoulet A, Lynch K, Wilson MR, Kämpe O, Dobbs K, Delmonte OM, Bacchetta R, Notarangelo LD, Burns JC, Casanova JL, Lionakis MS, Torgerson TR, Anderson MS, DeRisi JL. Autoantibody discovery across monogenic, acquired, and COVID-19-associated autoimmunity with scalable PhIP-seq. Rosen A, Rath S, Pillai S, editors. eLife. eLife Sciences Publications, Ltd; 2022 Oct 27;11:e78550.

16. Vazquez SE, Ferré EM, Scheel DW, Sunshine S, Miao B, Mandel-Brehm C, Quandt Z, Chan AY, Cheng M, German M, Lionakis M, DeRisi JL, Anderson MS. Identification of novel, clinically correlated autoantigens in the monogenic autoimmune syndrome APS1 by proteome-wide PhIP-Seq. Rosen A, Rath S, Craft J, editors. eLife. eLife Sciences Publications, Ltd; 2020 May 15;9:e55053.

17. Kelch-like Protein 11 Antibodies in Seminoma-Associated Paraneoplastic Encephalitis | NEJM [Internet]. [cited 2023 Jun 6]. Available from: https://www.nejm.org/doi/10.1056/NEJMoa1816721

18. O’Donovan B, Mandel-Brehm C, Vazquez SE, Liu J, Parent AV, Anderson MS, Kassimatis T, Zekeridou A, Hauser SL, Pittock SJ, Chow E, Wilson MR, DeRisi JL. High-resolution epitope mapping of anti-Hu and anti-Yo autoimmunity by programmable phage display. Brain Commun. 2020 Aug 3;2(2):fcaa059. PMCID: PMC7425417

19. Rackaityte E, Proekt I, Miller HS, Ramesh A, Brooks JF, Kung AF, Mandel-Brehm C, Yu D, Zamecnik C, Bair R, Vazquez SE, Sunshine S, Abram CL, Lowell CA, Rizzuto G, Wilson MR, Zikherman J, Anderson MS, DeRisi JL. Validation of a murine proteome-wide phage display library for the identification of autoantibody specificities [Internet]. bioRxiv; 2023 [cited 2023 Jun 10]. p. 2023.04.07.535899. Available from: https://www.biorxiv.org/content/10.1101/2023.04.07.535899v1

20. Bodansky A, Sabatino JJ, Vazquez SE, Chou J, Novak T, Moffitt KL, Miller HS, Kung AF, Rackaityte E, Zamecnik CR, Rajan JV, Kortbawi H, Mandel-Brehm C, Mitchell A, Wang CY, Saxena A, Zorn K, Yu DJL, Asaki J, Pluvinage JV, Wilson MR, Loftis LL, Hobbs CV, Tarquinio KM, Kong M, Fitzgerald JC, Espinal PS, Walker TC, Schwartz SP, Crandall H, Irby K, Staat MA, Rowan CM, Schuster JE, Halasa NB, Gertz SJ, Mack EH, Maddux AB, Cvijanovich NZ, Zinter MS, Zambrano LD, Campbell AP, Randolph AG, Anderson MS, DeRisi JL, Investigators the OC 19 NSG. A distinct cross-reactive autoimmune response in multisystem inflammatory syndrome in children (MIS-C) [Internet]. medRxiv; 2023 [cited 2023 Jun 26]. p. 2023.05.26.23290373. Available from: https://www.medrxiv.org/content/10.1101/2023.05.26.23290373v1

21. ZSCAN1 Autoantibodies Are Associated with Pediatric Paraneoplastic ROHHAD - Mandel-Brehm - 2022 - Annals of Neurology - Wiley Online Library [Internet]. [cited 2023 Jun 26]. Available from: https://onlinelibrary.wiley.com/doi/full/10.1002/ana.26380

22. Lohse AW, Hennes E. Diagnostic criteria for autoimmune hepatitis. Hepatol Res. 2007 Oct;37 Suppl 3:S509. PMID: 17931212

23. Taubert R, Engel B, Diestelhorst J, Hupa-Breier KL, Behrendt P, Baerlecken NT, Sühs KW, Janik MK, Zachou K, Sebode M, Schramm C, Londoño MC, Habes S, Consortium the UA, Oo YH, Lalanne C, Pape S, Schubert M, Hust M, Dübel S, Thevis M, Jonigk D, Beimdiek J, Buettner FFR, Drenth JPH, Muratori L, Adams DH, Dyson JK, Renand A, Graupera I, Lohse AW, Dalekos GN, Milkiewicz P, Stangel M, Maasoumy B, Witte T, Wedemeyer H, Manns MP, Jaeckel E. Quantification of polyreactive immunoglobulin G facilitates the diagnosis of autoimmune hepatitis. Hepatology. 2022;75(1):13–27.

24. Volkmann M, Luithle D, Zentgraf H, Schnölzer M, Fiedler S, Heid H, Schulze-Bergkamen A, Strassburg CP, Gehrke SG, Manns MP. SLA/LP/tRNP(Ser)Sec antigen in autoimmune hepatitis: Identification of the native protein in human hepatic cell extract. Journal of Autoimmunity. 2010 Feb 1;34(1):59–65.

25. Ezhilarasan D. Relaxin in hepatic fibrosis: What is known and where to head? Biochimie. 2021 Aug 1;187:144–151.

26. Hu M, Wang Y, Liu Z, Yu Z, Guan K, Liu M, Wang M, Tan J, Huang L. Hepatic macrophages act as a central hub for relaxin-mediated alleviation of liver fibrosis. Nat Nanotechnol. 2021 Apr;16(4):466–477. PMID: 33495618

27. Poetranto AL, Wakata A, Tjan LH, Nishimura M, Arii J, Mori Y. Human herpesvirus 6A U27 plays an essential role for the virus propagation. Microbiol Immunol. 2020 Oct;64(10):703– 711. PMID: 32827324

28. Herkel J, Heidrich B, Nieraad N, Wies I, Rother M, Lohse AW. Fine specificity of autoantibodies to soluble liver antigen and liver/pancreas. Hepatology. 2002;35(2):403– 408.

29. HHV-6 in liver transplantation: A literature review - Phan - 2018 - Liver International - Wiley Online Library [Internet]. [cited 2023 Jun 11]. Available from: https://onlinelibrary.wiley.com/doi/10.1111/liv.13506

30. Schmitt K, Deutsch J, Tulzer G, Meindl R, Aberle S. Autoimmune hepatitis and adrenal insufficiency in an infant with human herpesvirus-6 infection. The Lancet. Elsevier; 1996 Oct 5;348(9032):966. PMID: 8843841

31. Iredale JP, Pellicoro A, Fallowfield JA. Liver Fibrosis: Understanding the Dynamics of Bidirectional Wound Repair to Inform the Design of Markers and Therapies. Digestive Diseases. 2017 May 3;35(4):310–313.

32. Snowdon VK, Lachlan NJ, Hoy AM, Hadoke PWF, Semple SI, Patel D, Mungall W, Kendall TJ, Thomson A, Lennen RJ, Jansen MA, Moran CM, Pellicoro A, Ramachandran P, Shaw I, Aucott RL, Severin T, Saini R, Pak J, Yates D, Dongre N, Duffield JS, Webb DJ, Iredale JP, Hayes PC, Fallowfield JA. Serelaxin as a potential treatment for renal dysfunction in cirrhosis: Preclinical evaluation and results of a randomized phase 2 trial. PLoS Med. 2017 Feb;14(2):e1002248. PMCID: PMC5330452

33. Pettersen EF, Goddard TD, Huang CC, Meng EC, Couch GS, Croll TI, Morris JH, Ferrin TE. UCSF ChimeraX: Structure visualization for researchers, educators, and developers. Protein Sci. 2021 Jan;30(1):70–82. PMCID: PMC7737788

34. Petrie EJ, Lagaida S, Sethi A, Bathgate RAD, Gooley PR. In a Class of Their Own – RXFP1 and RXFP2 are Unique Members of the LGR Family. Frontiers in Endocrinology [Internet]. 2015 [cited 2023 Jun 11];6. Available from: https://www.frontiersin.org/articles/10.3389/fendo.2015.00137

35. Sethi A, Bruell S, Ryan T, Yan F, Tanipour MH, Mok YF, Draper-Joyce C, Khandokar Y, Metcalfe RD, Griffin MDW, Scott DJ, Hossain MA, Petrie EJ, Bathgate RAD, Gooley PR. Structural Insights into the Unique Modes of Relaxin-Binding and Tethered-Agonist Mediated Activation of RXFP1 and RXFP2. J Mol Biol. 2021 Oct 15;433(21):167217. PMID: 34454945

36. Parsell DA, Mak JY, Amento EP, Unemori EN. Relaxin Binds to and Elicits a Response from Cells of the Human Monocytic Cell Line, THP-1*. Journal of Biological Chemistry. 1996 Nov 1;271(44):27936–27941.

37. Hahn JW, Yang HR, Moon JS, Chang JY, Lee K, Kim GA, Rahmati M, Koyanagi A, Smith L, Kim MS, Sánchez GFL, Elena D, Shin JY, Shin JI, Kwon R, Kim S, Kim HJ, Lee H, Ko JS, Yon DK. Global incidence and prevalence of autoimmune hepatitis, 1970–2022: a systematic review and meta-analysis. eClinicalMedicine. 2023 Oct 17;65:102280. PMID: 37876996

38. Christen U, Hintermann E. Pathogen Infection as a Possible Cause for Autoimmune Hepatitis. International Reviews of Immunology. Taylor & Francis; 2014 Jul 4;33(4):296–313. PMID: 24911790

39. Christen U, Hintermann E. Animal Models for Autoimmune Hepatitis: Are Current Models Good Enough? Frontiers in Immunology. 2022 Jul 12;13:898615. PMID: 35903109

40. Adao I, Klepper A, Tana M. AIH Therapy: Beyond First-Line. Curr Hepatology Rep [Internet]. 2024 Mar 8 [cited 2024 Jul 22]; Available from: 10.1007/s11901-024-00657-4

41. Mitra A, Ray S. High-Titre ANA Positivity in MASLD: An Uncommon Presentation of a Common Disease. Eur J Case Rep Intern Med. 2020 Jun 8;7(9):001714. PMCID: PMC7473677

42. Saxinger C, Polesky H, Eby N, Grufferman S, Murphy R, Tegtmeir G, Parekh V, Memon S, Hung C. Antibody reactivity with HBLV (HHV-6) in U.S. populations. Journal of Virological Methods. 1988 Sep 1;21(1):199–208.

43. Bodansky A, Mettelman RC, Sabatino JJ, Vazquez SE, Chou J, Novak T, Moffitt KL, Miller HS, Kung AF, Rackaityte E, Zamecnik CR, Rajan JV, Kortbawi H, Mandel-Brehm C, Mitchell A, Wang CY, Saxena A, Zorn K, Yu DJL, Pogorelyy MV, Awad W, Kirk AM, Asaki J, Pluvinage JV, Wilson MR, Zambrano LD, Campbell AP, Thomas PG, Randolph AG, Anderson MS, DeRisi JL. Molecular mimicry in multisystem inflammatory syndrome in children. Nature. Nature Publishing Group; 2024 Aug;632(8025):622–629.

44. Zamecnik CR, Rajan JV, Yamauchi KA, Mann SA, Loudermilk RP, Sowa GM, Zorn KC, Alvarenga BD, Gaebler C, Caskey M, Stone M, Norris PJ, Gu W, Chiu CY, Ng D, Byrnes JR, Zhou XX, Wells JA, Robbiani DF, Nussenzweig MC, DeRisi JL, Wilson MR. ReScan, a Multiplex Diagnostic Pipeline, Pans Human Sera for SARS-CoV-2 Antigens. Cell Rep Med. 2020 Sep 24;1(7):100123. PMCID: PMC7513813

45. Pedregosa F, Varoquaux G, Gramfort A, Michel V, Thirion B, Grisel O, Blondel M, Prettenhofer P, Weiss R, Dubourg V, Vanderplas J, Passos A, Cournapeau D. Scikit-learn: Machine Learning in Python. MACHINE LEARNING IN PYTHON.

46. Vita R, Mahajan S, Overton JA, Dhanda SK, Martini S, Cantrell JR, Wheeler DK, Sette A, Peters B. The Immune Epitope Database (IEDB): 2018 update. Nucleic Acids Res. 2019 Jan 8;47(Database issue):D339–D343. PMCID: PMC6324067

